# Long-term course of humoral and cellular immune responses in outpatients after SARS-CoV-2 infection

**DOI:** 10.1101/2021.06.24.21259218

**Authors:** Julia Schiffner, Insa Backhaus, Jens Rimmele, Sören Schulz, Till Möhlenkamp, Julia Maria Klemens, Dorinja Zapf, Werner Solbach, Alexander Mischnik

**Affiliations:** University of Luebeck, Center for Infection and Inflammation Research, Luebeck, Germany; German Center for Infection Research (DZIF), Standort Hamburg-Borstel-Luebeck-Riems; Health Protection Authority, Luebeck, Germany; Institute of Medical Sociology, Centre for Health and Society, Medical Faculty and University Hospital, Heinrich-Heine-University, Düsseldorf, Germany; Institute for Experimental Immunology, affiliated to EUROIMMUN Medizinische Labordiagnostika AG, Luebeck, Germany

**Keywords:** COVID-19, SARS-CoV-2, immunoglobulin, IgG, seroprevalence, interferon-gamma release assay, IGRA

## Abstract

Characterisation of the naturally acquired B and T cell immune responses to SARS-CoV-2 is important for the development of public health and vaccination strategies to manage the burden of COVID-19 disease. We conducted a prospective, longitudinal analysis in COVID-19 recovered patients at various time points over a 10-month period in order to determine how circulating antibody levels and interferon-gamma (IFN-γ) release by peripheral blood cells change over time following natural infection.

From March 2020 till January 2021, we enrolled 412 adults mostly with mild or moderate disease course. At each study visit, subjects donated peripheral blood for testing of anti-SARS-CoV-2 IgG antibodies and IFN-γ release after SARS-CoV-2 S-protein stimulation. Anti-SARS-CoV-2 IgG antibodies were identified in 316/412 (76.7%) of the patients and 215/412 (52.2%) had positive neutralizing antibody levels. Likewise, in 274/412 (66.5 %) positive IFN-γ release and IgG antibodies were detected. With respect to time after infection, both IgG antibody levels and IFN-γ concentrations decreased by about half within three hundred days. Statistically, IgG and IFN-γ production were closely associated, but on an individual basis we observed patients with high antibody titres but low IFN-γ levels and vice versa.

Our data suggest that immunological reaction is acquired in most individuals after infection with SARS-CoV-2 and is sustained in the majority of patients for at least 10 months after infection. Since no robust marker for protection against COVID-19 exists so far, we recommend utilizing both, IgG and IFN-γ release for an individual assessment of immunity status.

## Introduction

Infection with severe acute respiratory syndrome coronavirus 2 (SARS-CoV-2) leads to various symptoms, including cough, fever, cold, and loss of smell and taste. The course of the disease varies in symptoms and severity, from asymptomatic infections to severe pneumonia with lung failure and death. Manifestation indices are estimated to be 55-85%^1^. About 48 % of patients are women, and 52 % are men. In Germany, 2.6 % of all persons with confirmed SARS-CoV-2 infections died in connection with a COVID-19 illness. The main risk factors for death are age and comorbidities like diabetes or obesity. The diagnosis is based on clinical grounds and proven by virus detection through RT-PCR in respiratory samples.

SARS-CoV-2 infects human cells by using the viral spike (S) protein, which binds to the angiotensin converting enzyme-2 (ACE-2) receptor on host cells^2^. The S-Protein is the immunodominant epitope that induces B and T-cell responses upon natural infection^3, 4^ and vaccination^5^. Antibodies target the virus and can block infection and, thus, are an essential correlate of protection^6, 7^. Likewise, T-lymphocytes contribute to protection through specific interactions with B-cells and cytokine responses^8^.

In the present study, we analysed the long-term course of the immune response with respect to serum IgG antibodies and the capacity of peripheral blood cells to produce interferon-gamma (IFN-γ) upon viral S-Protein specific stimulation. Based on previous experience of our group^9^ with the low diagnostic significance of IgA and IgM antibodies in the long-term course of infection, we deliberately determined only IgG antibodies.

The study was performed from May 2020 until January 2021. None of the study participants had received a COVID-19 vaccine.

## Materials and Methods

### Study area

The study was performed on patients which were notified as index cases to the Health Protection Authority of the City of Luebeck/Germany (approx. 220,000 inhabitants, population density approx. 1,000/sqm). With the exception of two major outbreak-related periods in December 2020 and mid-January 2021 the city has mostly been a low-incidence region, when compared to Germany as a whole (Fig. 1).

**Figure 1.:**
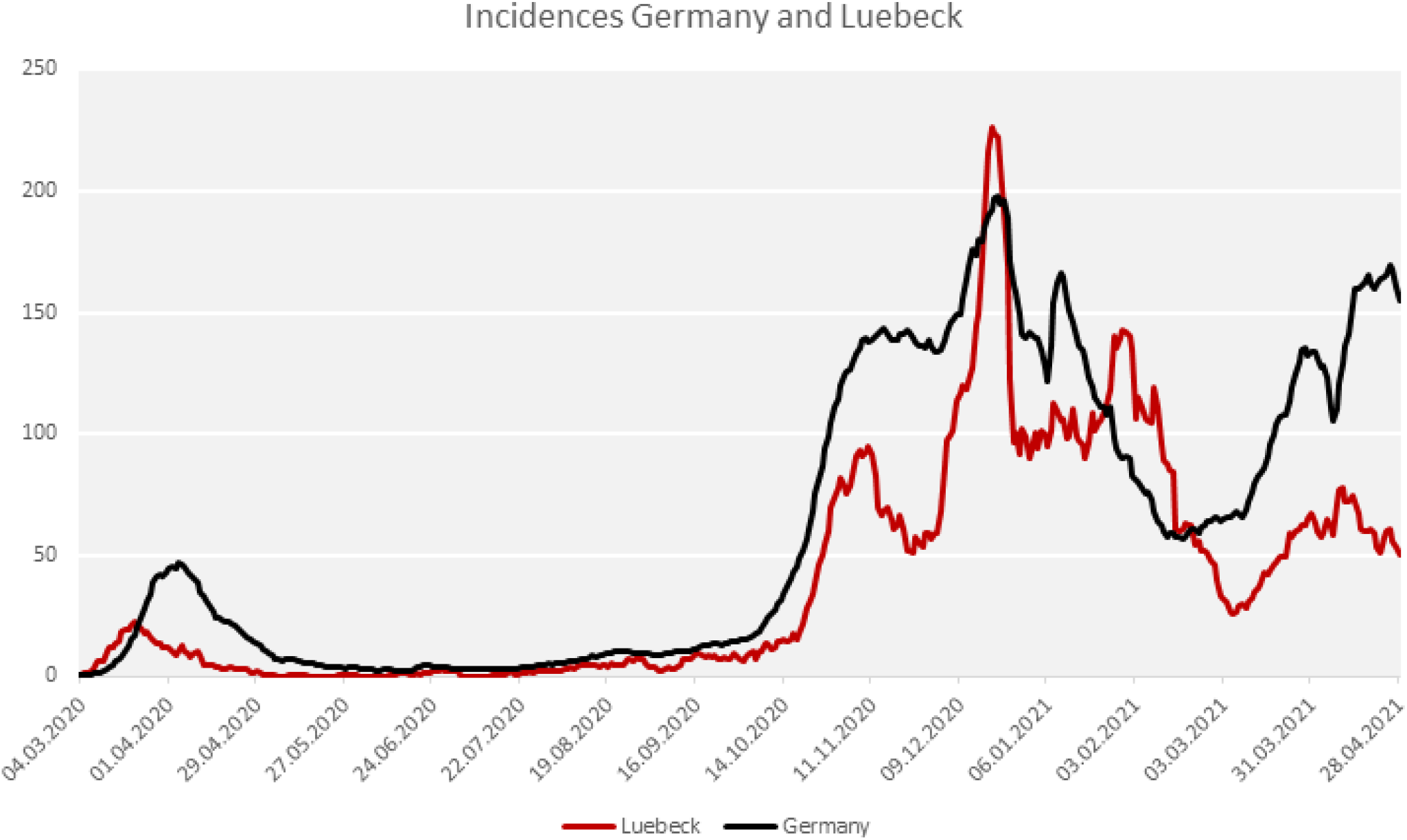
COVID-19-incidence in the city of Luebeck in comparison to Germany.

### Study population

The data presented here were obtained from the sera of patients that were notified to the local Health Protection Authority as being SARS-CoV-2 positive by PCR irrespective of the clinical manifestation. All of them recovered from the disease without hospitalization. In total, 1,279 patients were invited by e-mail to participate. 111 of these were first infected in the “first wave” of the pandemic between February 27, and July 31, 2020. The results of the initial antibody profiling from these patients have been published elsewhere.^15^ The remaining 1,168 patients that were invited were diagnosed by SARS-CoV-2 PCR between August 1, 2020 and December 31, 2021. From the invited patients, 436 responded to the invitation and, finally, 412 of them donated blood for analysis after having given written informed consent (see flowchart).

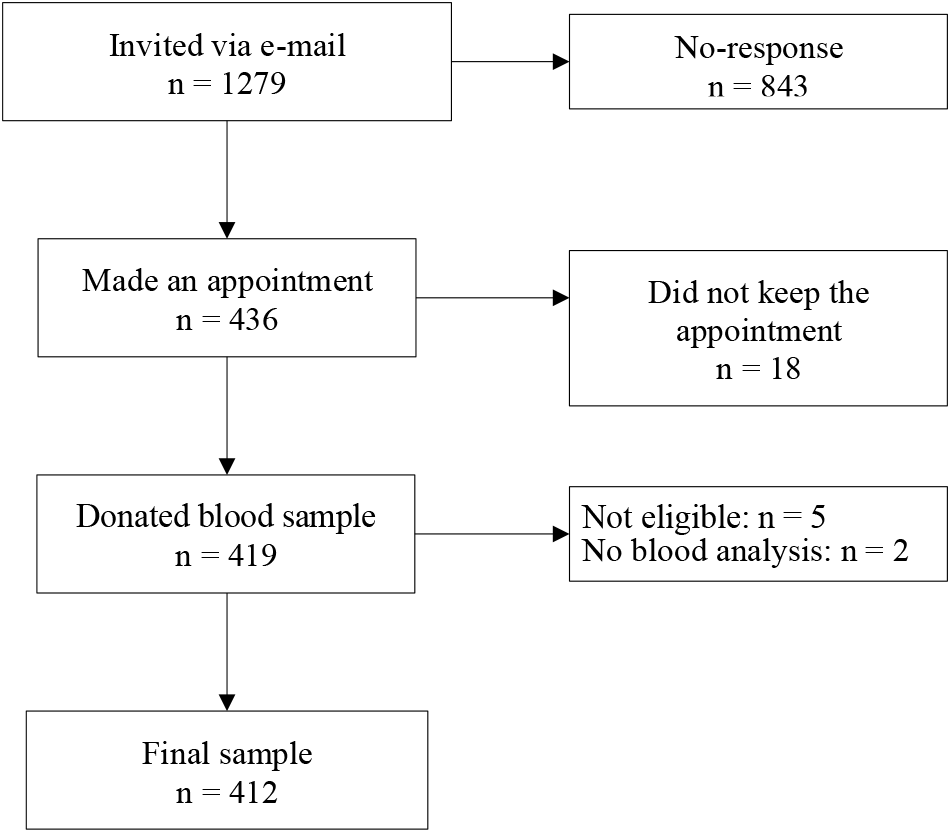

At the study visit, a questionnaire was filled in to determine clinical comorbidities (such as obesity, diabetes, autoimmune diseases, hypertonus).

COVID-19 disease severity was categorized based on the RKI severity definitions (RKI: klinische Klassifikation der COVID-19 Infektion adaptiert nach WHO Therapeutics and COVID-19: living guideline,https://www.rki.de/DE/Content/Kommissionen/Stakob/Stellungnahmen/Stellungnahme-Covid-19_Therapie_Diagnose.pdf?blob=publicationFile) and an additional category for patients that did not experience any symptoms of COVID-19 during the infection:

– asymptomatic
– mild (absence of pneumonia)
– moderate (signs of non-severe pneumonia)
– severe (severe pneumonia, defined as fever and bilateral pulmonary infiltrates and either respiratory rate> 30/min, severe respiratory distress or SpO2 < 90-94 % on room air)
– critical (acute respiratory distress syndrome; hyperinflammation in conjunction with sepsis or septic shock and multiple organ failure)

### Test procedures

#### Detection of SARS-CoV-2

Nasopharyngeal swabs were taken from suspected COVID-19 cases by trained personnel either in general practice (GP) or in a “drive-in” swab centre run by the Health Protection Authority between March 2020 and December 2020. Swabs were stored in stabilization media and laboratory-processed within 4 hours. SARS-CoV-2 RNA was detected by using an automated one step real-time rt-PCR (RIDA^®^GENE SARS-CoV-2 RUO Test; R-Biopharm AG, Darmstadt, Germany; E-gene amplification) run on a RIDA^®^CYCLER according to the manufacturer’s instruction.

#### Detection of anti-SARS-CoV-2 S1-Protein IgG antibodies

Serum Anti-SARS-CoV-2 IgG were detected by automated Enzyme-linked Immunosorbent Assay (product EI 2606-9601 G; EUROIMMUN; https://www.euroimmun.com) according to the manufacturer’s instructions. Signal-to-cut-off (SCO) ratio was calculated as extinction value (450 nm) of patient sample divided by extinction level of calibrator. A ratio between 0 and <0.8 was considered as negative, ≥ 0.8 to < 1.1 as borderline and a ratio ≥ 1.1 as positive. Assay specificity using pre-COVID-19 samples was calculated by the manufacturer as 100 %.

#### Detection of neutralizing antibodies

For detection of neutralizing antibodies, a semi-quantitative surrogate virus neutralization test (NeutraLISA from Euroimmun, Product No. 2606-4) was applied, through which the binding of SARS-CoV-2 S1/receptor-binding-domain RBD to ACE2 receptors of the recombinant human host cells is determined. In the first reaction step, samples and controls are incubated with soluble biotinylated ACE. If neutralizing antibodies are present in the sample, they compete with the ACE-receptor for the binding site of the SARS-CoV-2 S1/RBD proteins. Unbound ACE is removed through washing. To detect the bound ACE, a second incubation step with peroxidase-labeled streptavidin is performed, which catalyzes a color reaction. The intensity of the formed color is inversely proportional to the concentration of neutralizing antibodies in the sample. The inhibition (% IH) is calculated to the formular: % IH = 100-(extinction of patient sample x 100/extinction of blank). Values below 20 are considered negative, ≥ 20 to < 35 as borderline and ≥ 35 as positive. According to the manufacturer, sensitivity and specificity are calculated as 95.9% and 99.7%, respectively.

#### Detection of T-cell-activity

Besides B cells and antibodies, T lymphocytes and cytokines are instrumental for the shaping of the specific acute and memory immune response to SARS-CoV-2^10^. We sought for an easy-to-perform test that may be used as a marker for T cell responses by determining the capacity to release Interferon-gamma (IFN-γ) upon specific stimulation. 7 mL blood was collected in heparinized blood collection tubes. Within 6 hours 0.5 mL of the blood was transferred into three different tubes. One positive control tube (containing a mitogen), one SARS-CoV-2 specific stimulation tube (coated with antigens based on the SARS-CoV-2 spike protein) and one blank tube without antigens to measure one’s individual IFN-γ background (product ET 2606-3003, https://www.euroimmun.com). After 20 to 24 hours of incubation at 37 °C, the tubes were centrifuged at 12.000 rfc for 10 minutes. IFN-γ concentrations were measured in the supernatants by IFN-γ ELISA according to the manufacturer’s instructions (product EQ 6841-9601, EUROIMMUN; https://www.euroimmun.com). When the positive control tube shows a reaction (to confirm sufficient quantity and viability of immune cells) the IFN-γ concentration from the specific stimulation tube (after substracting the IFN-γ background) was used to quantify specific T cell responses. Values ≥ 100 mIU/mL were interpreted as borderline, ≥ 200 mIU/mL as positive. It has to be mentioned that all IFN-γ concentrations above the measurement range were replaced by the numerical value of 2500 mIU/ml. A further quantification was not possible.

#### Statistical analysis

We used standard descriptive statistics to summarize data. Categorical data were presented as frequencies and percentages and continuous variables were expressed as the mean with ± standard deviation (SD). Categorical variables were compared using Fisher’s exact test and continuous variables were compared using the Mann-Whitney U test. Pearson’s correlation coefficient (Pearson’s *R)* and p-value were calculated to evaluate the correlation between variables, where a Person’s *R* between 0-0.19 is regarded as very weak, 0.2-0.39 as weak, 0.40-0.59 as moderate, 0.6-0.79 as strong and 0.8-1 as very strong correlation^11^. To identify potential factors associated with SARS-CoV-2 seropositivity (yes/no), a binary logistic regression analysis using the logitem command in Stata (StataCorp, College Station, TX, version 15) to correct for the test’s specificity and sensitivity was performed. We estimated the models with the following explanatory variables: comorbidity, COVID-19 disease course and time since the positive COVID-19 test by rt-PCR. Two separate models were run to account for potential confounding. Model 1 included an unadjusted analysis and Model 2 included age and sex-adjusted analysis. Confounding occurs in epidemiological research when the relationship between a given exposure and a specific outcome (i.e., seropositivity) is distorted (confused) by the influence of a third variable or group of variables (confounders). In the present analysis, age and sex were considered confounders if they changed the coefficient of the significant variables by >10%. 95% confidence intervals for odds ratios were calculated and a p-value of 0.05 was considered significant. Statistical analyses using were conducted using Stata version 15.0.

## Results

### Sociodemographic characteristics of the study participants

The age of the patients was between 16 and 83 years with a mean age of 44.5 years (SD: ± 16); (Fig. 2a). 235 patients (57 %) were female and 177 (43 %) were male (Fig. 2b). Approx. 40 % of the patients reported at least one comorbidity such as obesity, diabetes, autoimmune diseases or hypertonus (Fig. 2c). Around 90 % of the participants reported symptoms during the infection time, 8.9 % had no symptoms. About half of the patients were classified as having had mild disease, 36 % as moderate and only 15 patients (3.6 %) had severe disease, but no requirement for hospitalization (Fig. 2d). At the study time in January 2021, 41.5 % of the patients about three months after infection reported still to have everyday life restricting symptoms such as fatigue (21 %), disturbance of smell and/or taste (12.5%) and lack of concentration (8 %). Nearly 13 % of respondents described more than one persisting symptom (data not shown).

**Figure 2.:**
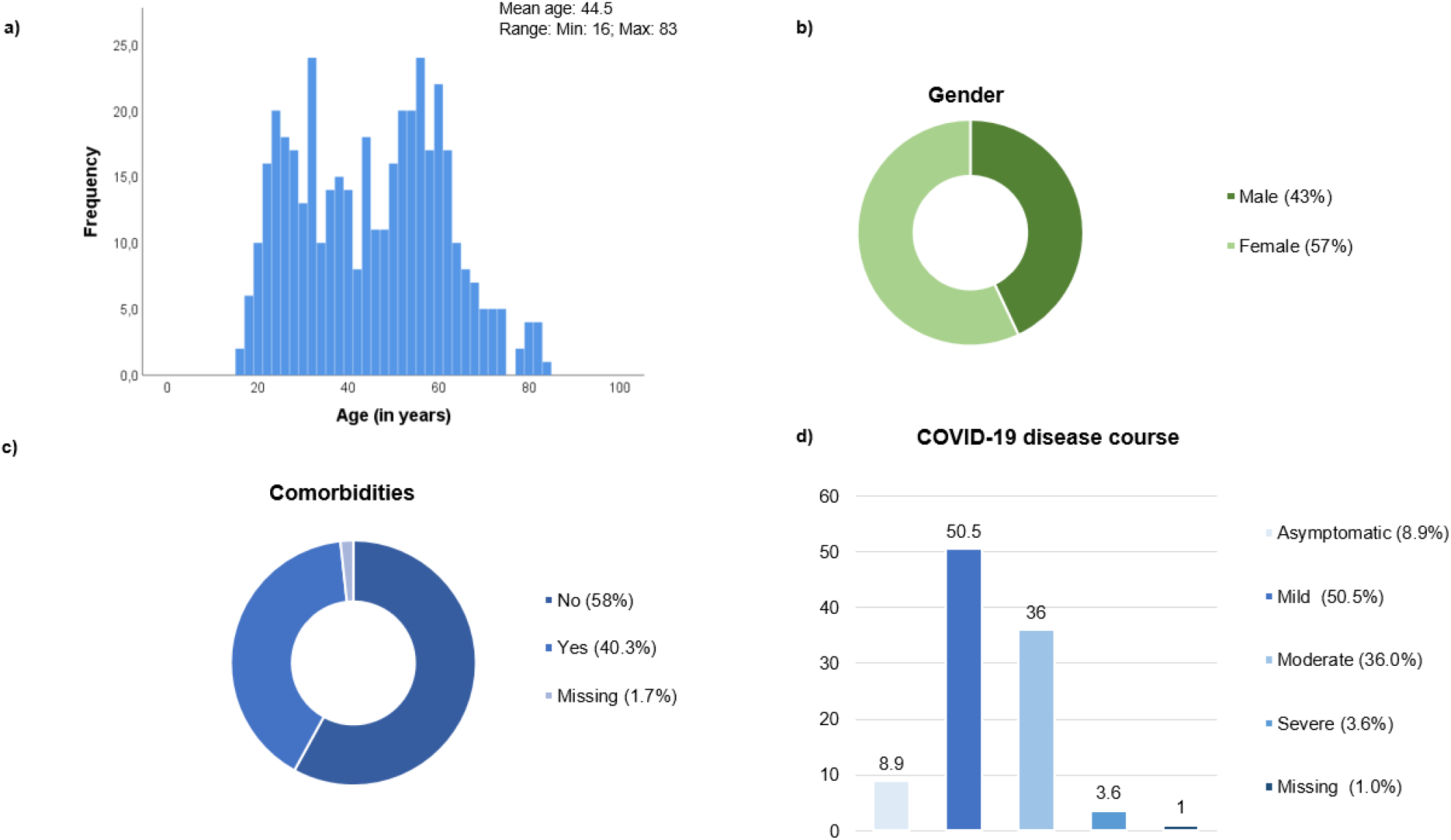
Sociodemographic characteristics of the study participants (n=412)

### IgG antibodies over time

There was a wide inter-individual variation in the antibody levels, supporting the observation from our earlier study^9^. Seropositivity was detected 16 days after the confirmed SARS-CoV-2 diagnosis by PCR (Fig. 3). Given that antibody ratios ≥ 1.1 are defined as positive, it is noteworthy that 16 % (66/412) of the participants did not develop humoral antibodies, despite SARS-CoV-2 detection by PCR. Clinically, the seronegative patients were either asymptomatic, in category 1 (mild) or 2 (moderate). The antibodies had S-protein neutralizing capacity. Most of the sera from the 346 IgG-positive patients (ratio ≥ 1.1), had neutralizing capacity (Inhibition index ≥ 20) 215/346 (62.1%) participants were neutralizing antibody positive (Inhibition index ≥ 20) and 88 (25.4%) participants showed borderline results.

**Figure 3.:**
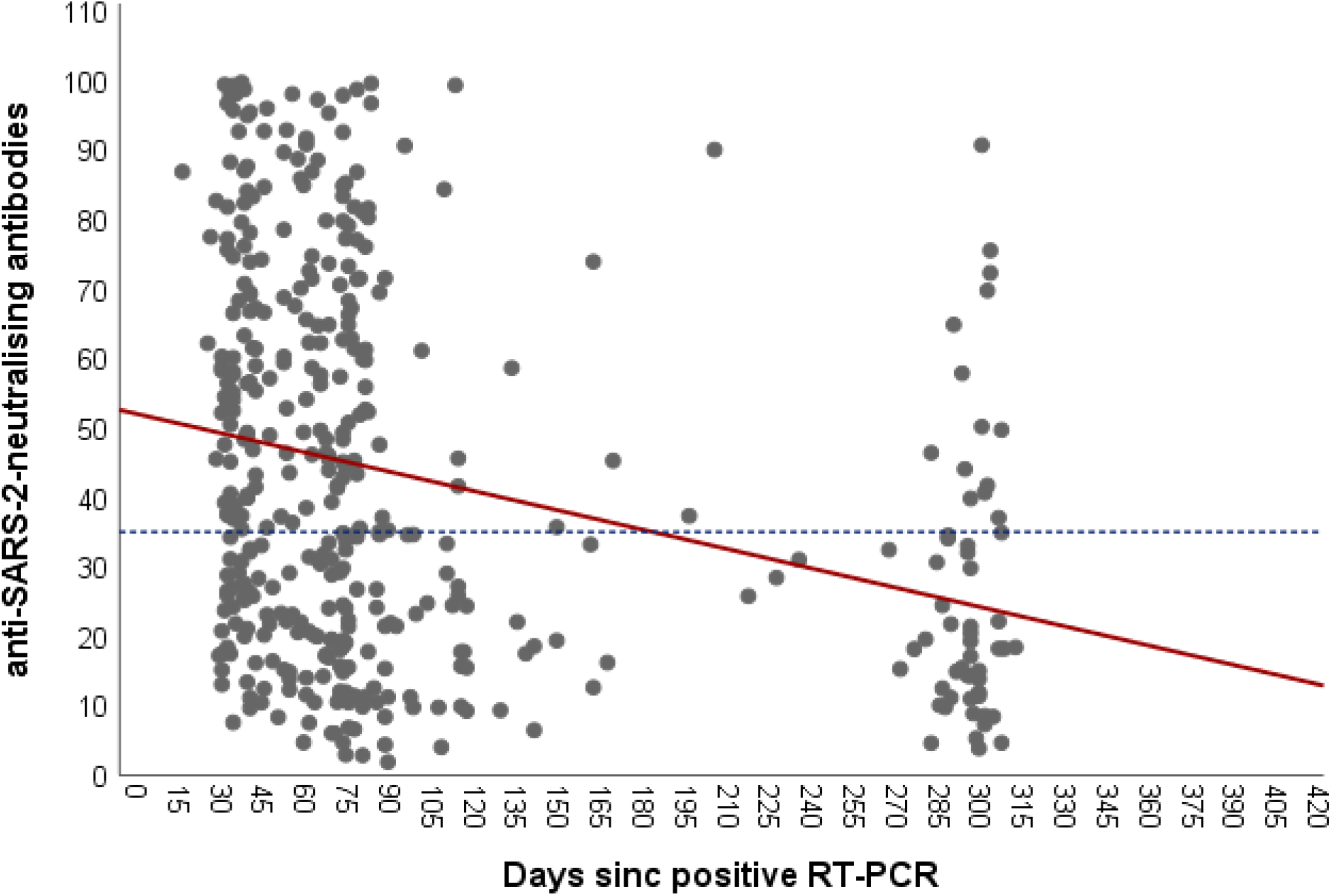
Correlation between anti-SARS-CoV-2-neutralising antibodies and time after COVID-19 infection. **Legend:** Scatter plot for anti-SARS-CoV-2-neutralising levels at different time points. Each black dot represents one participant and the anti-SARS-CoV-2-neutralising levels. The redline represents the interpretation line, which shows the negative linear association between the anti-SARS-CoV-2-neutralising level and the days passed since the COVID-19 diagnosis. The dotted line represents reference line for the cut-off for having anti-SARS-CoV-2-neutralising antibodies, which is set at 35.

Fig. 3 shows the antibody levels for IgG in relation to the days after PCR-positivity for the entire sample (n=412). It can be clearly seen that the antibody levels decline over time. There was a moderate but significant negative linear relationship between IgG ratio and the time passed since the positive SARS-CoV-2 diagnosis (r = -0.3, p < 0.001) (Fig. 3).

When we sought for factors that were associated with the presence of antibodies, it became clear that disease severity was positively associated, whereas reported comorbidities had no impact on antibody development (Model I; OR: 1.69, 95% CI: 1.10 – 2.84; Table 2). As could be seen in Fig. 3, we also found a significant association between the elapsed time of PCR testing and SARS-CoV-2 seropositivity. The likelihood to be seropositive weaned over time with an odds ratio (OR) of 0.38 in the time period of 60 - 119 days since PCR-positivity. The probability further decreased after 120 days to an OR of 0,19 (Model I; OR: 0.19, 95%CI: 0.09 – 0.37) (Table 2). After adjusting for age and sex as potential confounders, the variables (i.) moderate to severe disease course (Model II; OR: 1.77, 95% CI: 1.04 – 3.01) and (ii.) time of diagnosis remained significantly associated with being seropositive (Model II; OR: 0.18, 95%CI: 0.09 – 0.37). Furthermore, the estimates did not significantly change when adjusting for sex and age. Thus, in the present analysis both age and sex are not confounders. Although not statistically significant, a tendency for a greater odd of being seropositive were seen among participants having at least one comorbidity (Model II; OR: 1.07, 95% CI: 0.63 – 1.85) or were of advanced age (Model II; OR: 2.91, 95% CI:0.60 – 9.65). For female participants the odds ratio for being seropositive was almost equal to males (Model II; OR, 1.05, 95%CI: 0.64 – 1.73), suggesting that being seropositive is equally likely to occur in both female and male participants.

**Table 1.** Sample characteristics (n=412)

| **Table 1. Sample characteristics (n=412)** |  |
| --- | --- |
| **Characteristics** | **N (%)** |
| **Gender** |  |
| Male | 177 (43.0) |
| Female | 235 (57.0) |
| **Mean Age (±SD)** | 44.5 (16.0) |
| **Comorbidity** |  |
| Yes | 166 (40.3) |
| No | 239 (58.0) |
| Missing | 7 (1.7) |
| **COVID-19 disease course** |  |
| Asymptomatic | 36 (8.7) |
| Mild disease course | 209 (50.7) |
| Moderate disease course (fever, cough, trouble breathing) | 148 (35.9) |
| Severe disease course | 15 (3.6) |
| Missing | 4 (1.0) |
| **SARS-CoV-2 IgG antibodies** |  |
| Negative | 65 (15.8) |
| Borderline | 31 (7.5) |
| Positive | 316 (76.7) |
| **SARS-CoV-2 neutralizing antibodies** |  |
| Negative | 109 (26.5) |
| Borderline | 88 (21.4) |
| Positive | 215 (52.2) |
| **IGRA and IgG antibodies positive** | 274 (66.5) |
| **IGRA positive and IgG antibodies negative** | 46 (11.2) |
| **IgG antibodies positive and IGRA negative** | 42 (10.2) |

**Table 2.** Factors associated with the presence of SARS-CoV-2 IgG antibodies (n=412)

|  | **Model I**<br>**Crude OR [95% CI]** | **Model II**<br>**Adjusted OR [95% CI]** |
| --- | --- | --- |
| **Disease course** |  |  |
| Asymptomatic/Mild | 1.0 | 1.0 |
| Moderate/Severe | **1.69 [1.10 – 2.84]** | **1.77 [1.04 – 3.01]** |
| **Comorbidity** |  |  |
| Yes | 1.0 | 1.0 |
| No | 1.06 [0.64 – 1.75] | 1.07 [0.63 – 1.85] |
| **Time since positive RT-PCR** |  |  |
| 0-59 days | 1.0 | 1.0 |
| 60-119 days | **0.38 [0.21 – 0.69]** | **0.35 [0.19 – 0.65]** |
| 120 days and more | **0.19 [0.09 – 0.37]** | **0.18 [0.09 – 0.37]** |
| **Sex** |  |  |
| Male | - | 1.0 |
| Female | - | 1.05 [0.64 – 1.73] |
| **Age group** |  |  |
| 16-29 years | - | 1.0 |
| 30-49 years | - | 1.20 [0.63 – 2.32] |
| 50-69 years | - | 0.99 [0.44 – 1.64] |
| 70 years and older | - | 2.91 [0.60 – 9.65] |

**Legend:** 1.0 = reference category; OR: Odd Ratio; CI: Confidential Interval; Table 2 presents the unadjusted (Model I) and the age and sex-adjusted odds ratio (OR) (Model II) of seropositivity (Yes/No) by participant characteristics. The odds ratio was calculated to describe the risk of different groups in positive ELISA serums compared with non-positive ELISA sera. n = 412.

The odds of having antibodies with neutralizing capacity were almost two-fold higher in participants who had a moderate to severe COVID-19 disease course as compared to those with none or mild disease (Model I; OR. 1.96, 95% CI: 1.18 – 2.27; Table 3). Similar to IgG SARS-CoV-2 antibodies, elapsed time since the confirmed COVID-19 diagnosis was inversely associated with the probability of expressing neutralizing antibodies with a clear weaning after 60 to 119 days and even more after 120 days (Model I, OR. 0.15, 95% CI: 0.08 – 0.31). Individuals for which the time since SARS-CoV-2 testing was more than 120 days ago were less likely to have neutralizing antibodies (OR: 0.14; 95% CI: 0.07 – 0.28) compared to those with a diagnosis made between 0-59 days apart from the time of the serological survey. For female participants, although not statistically significant, the odds ratio for having neutralizing antibodies was slightly decreased (Model II; OR, 0:94; 95%CI: 0.58 – 1.52). The odds ratios in Model II did not significantly change when adjusting for age and sex, suggesting that both variables are not confounding the association between disease course, time since rt-PCR testing and having SARS-CoV-2 neutralizing antibodies.

**Table 3.** Factors associated with the presence of SARS-CoV-2 neutralizing antibodies (n=412)

|  | **Model I**<br>**Crude OR [95% CI]** | **Model II**<br>**Adjusted OR [95% CI]** |
| --- | --- | --- |
| **Disease course** |  |  |
| Asymptomatic/Mild | 1.0 | 1.0 |
| Moderate/Severe | **1.96 [1.18 – 2.27]** | **2.07 [1.23 - 3.47]** |
| **Comorbidity** |  |  |
| Yes | 1.0 | 1.0 |
| No | 1.39 [0.85 – 2.28] | 1.34 [0.79 – 2.27] |
| **Time since positive RT-PCR** |  |  |
| 0-59 days |  | 1.0 |
| 60-119 days | **0.39 [0.22 – 0.70]** | **0.35 [0.14 – 0.63]** |
| 120 days and more | **0.15 [0.08 – 0.31]** | **0.14 [0.07 – 0.28]** |
| **Sex** |  |  |
| Male |  | 1.0 |
| Female |  | 0.94 [0.58 – 1.52] |
| **Age group** |  |  |
| 16-29 years |  | 1.0 |
| 30-49 years |  | 1.02 [0.55 – 1.89] |
| 50-69 years |  | 0.93 [0.53 – 1.78] |
| 70 years and older |  | 2.56 [0.84 – 10.11] |

**Legend:** 1.0 = reference category; OR: Odd Ratio; CI: Confidential Interval; Table 3 presents the unadjusted (Model I) and the age and sex-adjusted odds ratio (OR) (Model II) of having neutralizing antibodies (Yes/No) by participant characteristics. The odds ratio was calculated to describe the factors of different groups in positive neutralizing antibody assays compared with negative neutralizing antibody assays.

The mean antibody ratio was highest in the first 1-3 months in patients with a severe disease course. In asymptomatic or mild symptomatic patients, the highest mean antibody level, although at a lower level, was also observed 1-3 months post COVID-19 infection, with a declining trend in the subsequent time windows (Table 4.) The data thus clearly show that antibody expression is related to the severity of disease and that antibody levels fade continuously within approx. 300 days (Fig. 3, Table 4).

**Table 4.** Mean antibody ratio since confirmed COVID-19 diagnosis and disease severity.

| Disease course | Days since confirmed COVID-19 diagnosis |  |
| --- | --- | --- |
|  | 1-3 months | 3 and more months |
| | Mean (±SD) | Mean (±SD) |
| Asymptomatic/ Mild | 3.38 (2.56) | 1.78 (1.53) |
| Moderate/Severe | 4.28 (2.22) | 2.26 (1.87) |

### T-cell-activity over time

Since T-cells are instrumental for the development of the S-protein reactive B cell activity, we determined T-lymphocyte activity. We chose to determine the induction and release of IFN-γ upon S-protein specific stimulation. This assay reflects an easy-to-perform summarized image of T-lymphocytes-activity without considering all subgroups of T-lymphocytes or other IFN-γ producing cells. As can be seen in Fig. 4, 315/412 (81 %) of the PCR-positive patients had a positive IGRA test result (≥ 200 mIU/ml). 62 (15 %) of the patients were negative. Like it was seen with the antibody analysis, there was strong relationship between the IFN-γ levels and the time elapsed since the positive SARS-CoV-2 diagnosis by PCR (r = 0.6, p < 0.001).

**Figure 4.:**
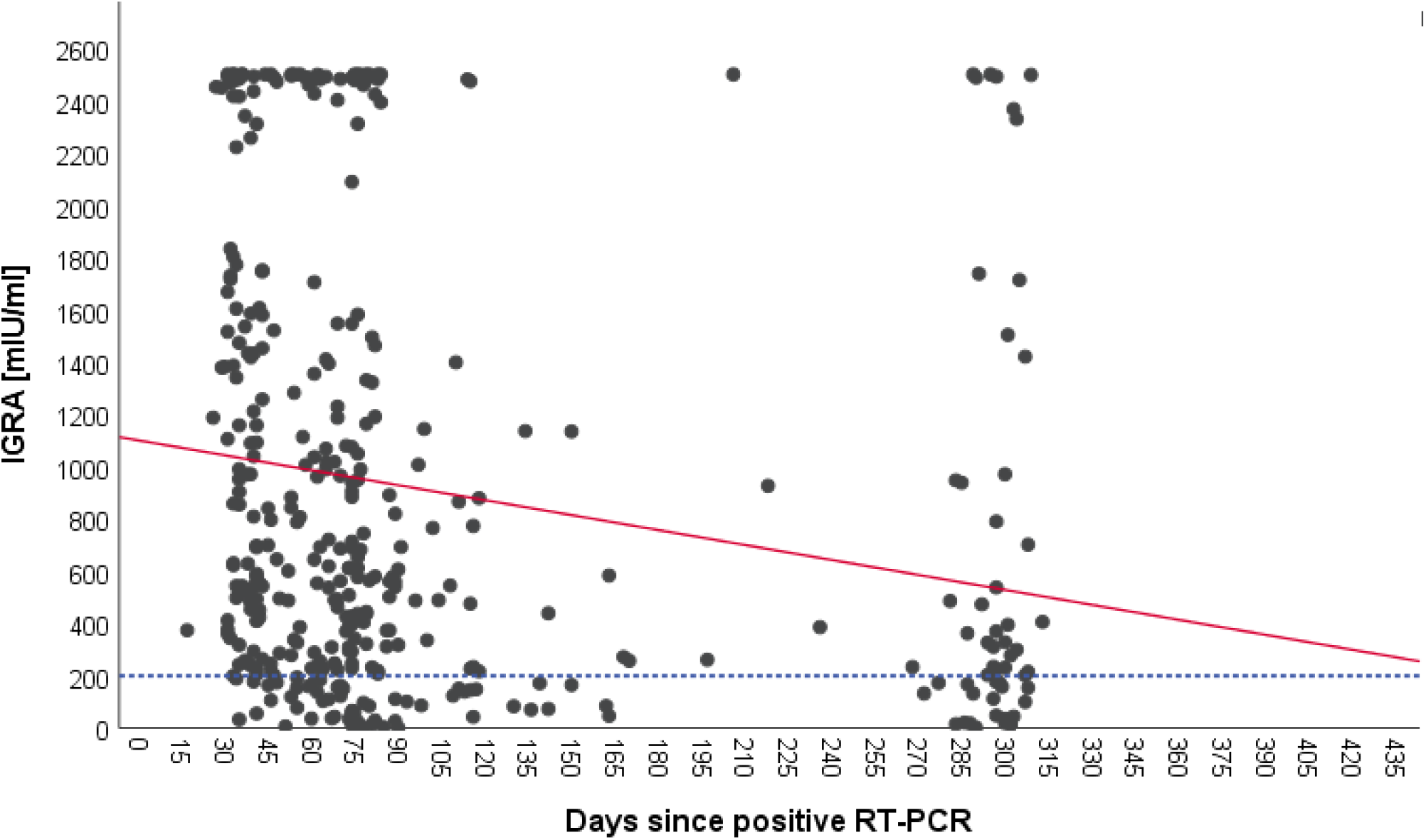
Correlation between IGRA and time after COVID-19 infection. **Legend:** Scatter plot for IGRA values at different time points. Each black dot represents one participant and the IGRA mIU/ml and the redline represent the interpretation line, which shows the negative linear association between IGRA values and the days passed since the COVID-19 diagnosis. The dotted line represents the cut-off of IGRA values at 200 mIU/ml.

Looking at the correlation between antibody levels and IFN-γ concentrations, a heterogeneous picture emerged (Fig. 5). While in most cases both values were concordant, there were a considerable number of cases with high to very high IFN-γ levels and low antibody levels and vice versa. Looking at the cloud of dots, it is striking that there appears to be a population of patients whose cells produce extremely high levels of IFN-γ (> 2,500 mIU/ml), regardless of the antibody response (Fig. 4) or the time post SARS-CoV-2 diagnosis (Fig. 3).

**Figure 5.:**
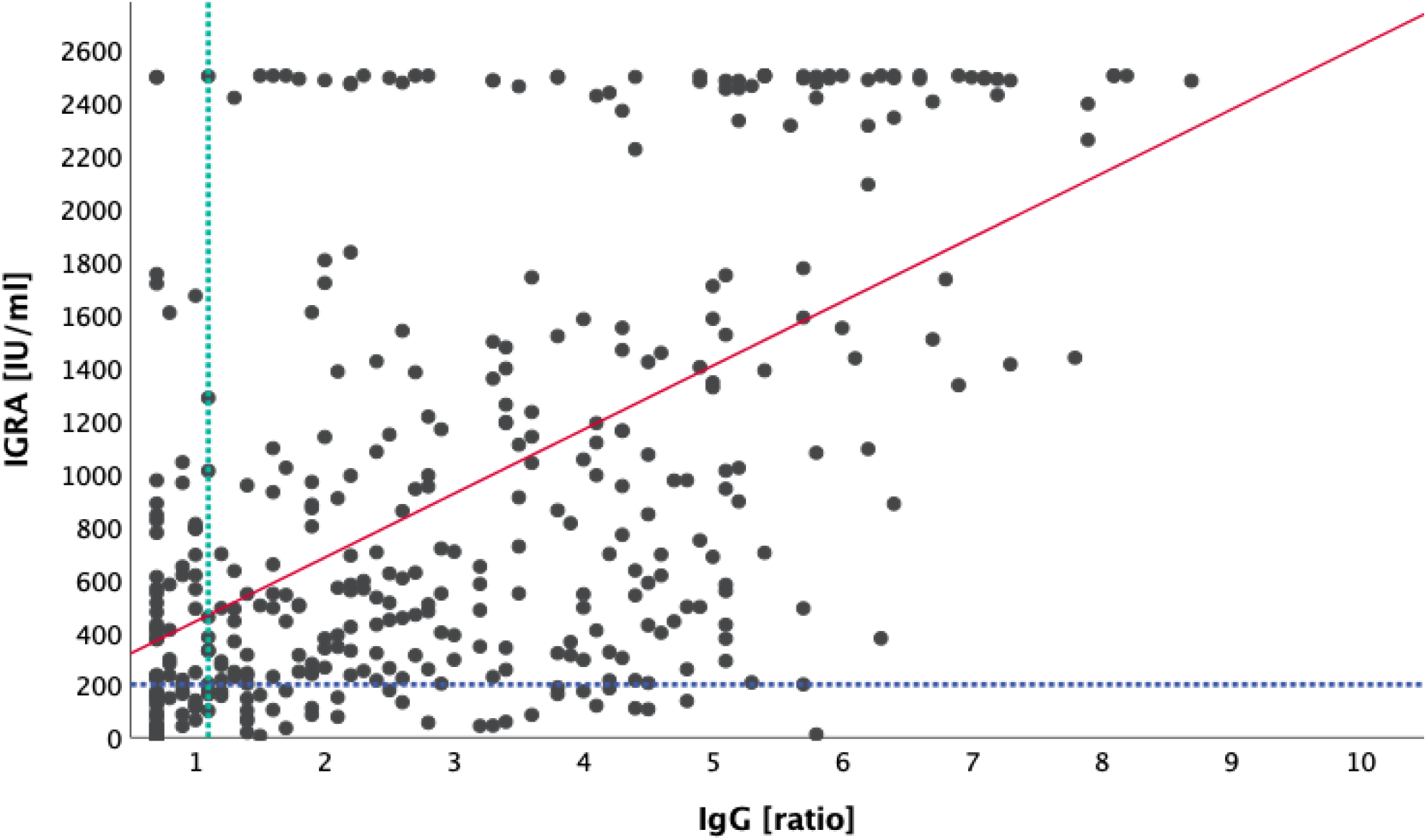
IgG antibodies versus IGRA. **Legend:** Scatter plot for IGRA values and IgG values. Each black dot represents one participant and the respective IgG and IGRA value. The dotted line represents the cut-off for positive IgG values at 1.1 (vertical line) and for IGRA values at 200 mIU/ml (horizontal line). The red line shows the significant positive correlation between IgG and IGRA values (r = 0.6, p < 0.001).

**Figure 6:**
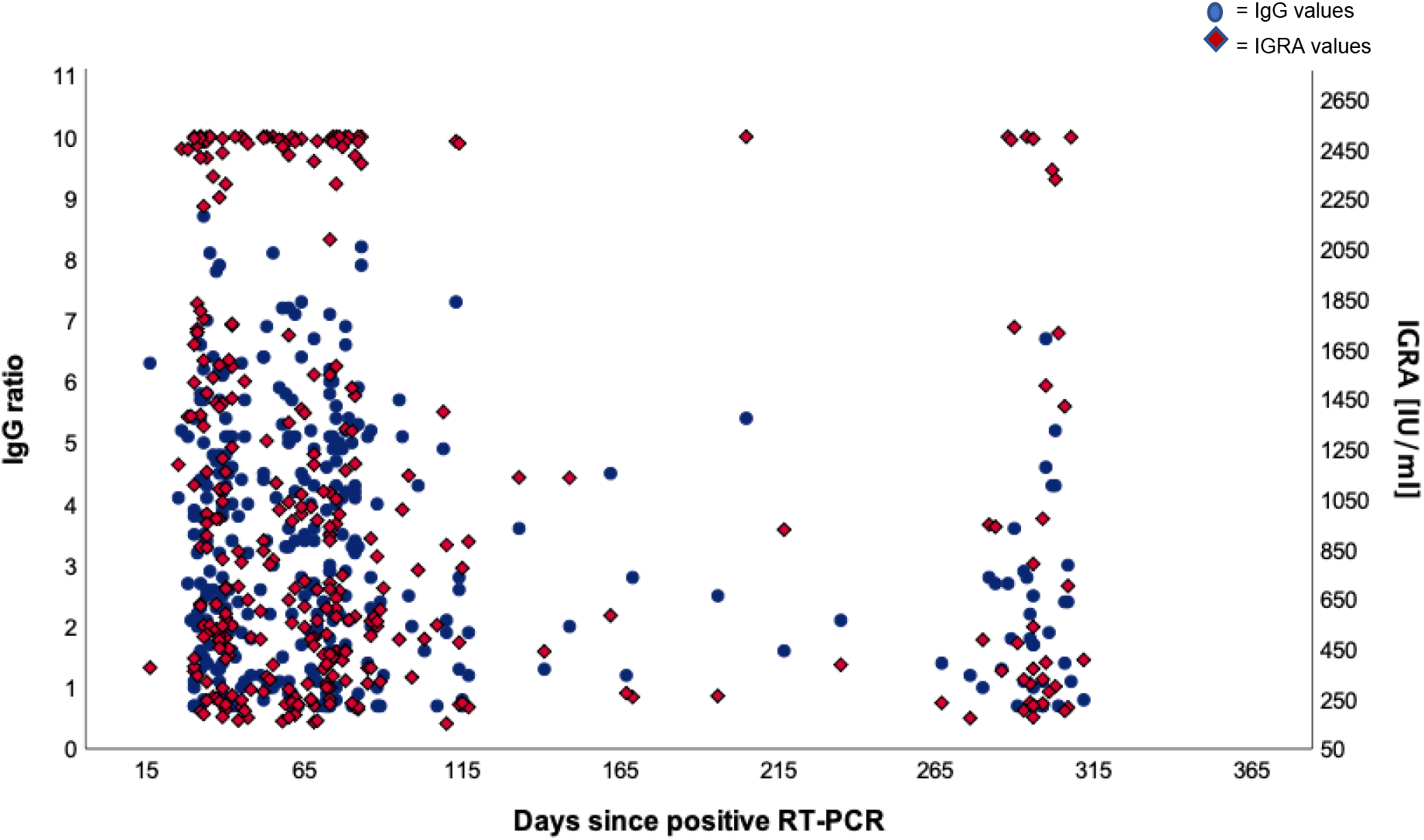
Dual axis scatter plot for IgG ratios and IGRA values. Legend: The dots represent participants and their positive IgG values (cut-off at 1.1) and the diamonds represent participants and their positive IGRA values (cut-off at 200 mIU/ml).

In summary, in our SARS-CoV-2 PCR-positive cohort, the antibody profile was heterogeneous. In most cases, antibodies could be detected between 16 and 73 days after the determination of a SARS-CoV-2 infection via PCR from a nasopharyngeal swab. The antibody levels continuously decreased over time. In 90/412 patients (22 %) no significant antibody levels could be detected in 2 up to 4 consecutive analyses between day 16 and 73. As far as the IFN-γ release is concerned, a similar picture emerged. The levels were heterogeneous and decreased over time. Noteworthy, a significant part of the patients produced very high levels of IFN-γ irrespective of the concentration of antibodies measured.

## Discussion

In the present study, we determined the development of specific humoral and cellular immune responses in outpatients recovered from SARS-CoV-2 infection, mostly with mild to moderate disease (Fig. 2). Therefore, we chose to determine serum IgG antibodies and IFN-γ release in response to the viral spike (S) glycoprotein in view of the time that had passed since the infection.

In the present study about 22 % of the patients had no detectable antibodies. These data substantiate our previous finding from the first wave of the pandemic in early 2020^9^. On a population basis, the clinical severity of the disease was positively correlated with the level of SARS-CoV-2 neutralising antibodies (Tables 3, 4), as had been shown previously by others^12^. On an individual level, there was a great variability between patients. Thus, the individual level of antibodies is not of diagnostic value, for example for the assessment of patients with long-COVID syndrome, which is often associated with chronic fatigue^13^. As expected, in many cases the antibody levels faded over time. When compared to the first three months after infection, the mean antibody levels decayed steadily and approximately halved within 300 days (Fig. 3). Although our data do not allow a meaningful calculation of the half-life, the finding is in accordance with reports from others^14, 15^, who calculated the half-life of neutralizing IgG antibodies with 140 to 220 days.

Interestingly, after COVID vaccination neutralisation capacity was different from natural infection^16^.

Very few published data sets compare antigen-specific B-cell and T-cell immunity. We therefore examined interrelationships between IgG antibody levels and IFN-γ release. Like with the antibody kinetics, the IFN-γ values decayed over time after infection with similar kinetics (Fig. 4) which is in line with other works^17^. Unexpectedly, however, we saw a substantial number of patients with low antibody levels and extremely high IFN-γ levels and vice versa (Fig. 5). Although unlikely, it cannot be completely ruled out, that the observed T-cell reactivity was due to pre-existing memory T-cells recognizing the common cold coronaviruses, as has been described before^18,19^. Existing T cells might be an explanation for cross reactions in an IGRA assay which could also be seen in our study^20^. To explain this dichotomy, a longitudinal in-depth analysis of the precise numbers and types of IFN-γ producing cells will be necessary, especially the characterization of T memory cells.

Since our data and those from others show that determination of antibodies alone is not predictive for protection against SARS-CoV-2 disease, simultaneous determination of IFN-γ may be a valuable adjunct and may also predict the time-point for possibly necessary booster vaccinations on an individual basis.

### Strengths and limitations

The current study has few limitations. Longitudinal data for each subject, with at least three time points per subject, would be required for more precise understanding of the kinetics of durability of SARS-CoV-2–specific antibodies. Nevertheless, the current cross-sectional data describe well the dynamics of spike-specific antibodies over 10 months and IFN-γ release by blood T-lymphocytes at one study point. This study was not sufficiently powered to control for many variables simultaneously.

## Conclusion

This study shows that on average 9.8 months after detection of SARS-CoV-2 in nasopharyngeal specimen, the mean serum IgG antibodies against the viral S-protein approximately dropped to 50 % of the initial values. Most of the patients showed robust IFN-γ production after S-protein stimulation of peripheral blood cells, indicating the importance of T lymphocytes for shaping the protective immune reaction. However, in a substantial proportion of our samples, we found low antibody levels accompanied with high IFN-γ levels and vice versa. For future assessment of protection and possible vaccination strategies, both, determination of antibodies and IFN-γ is recommended.

## Data Availability

All data are available if you contact the authors.

## Authors’ contributions

J.S., I.B., JR., A.M. and W.S. wrote the article. J.S., I.B., D.Z., J.M.K. and W.S. were responsible for data analysis. S.S., T.M. and J.R. were responsible for patient recruitment, data collection and contributed to data analysis. A.M. and W.S. designed the study. All authors approved the final version of the manuscript.

## Acknowledgment

The authors would like to thank all participants for sharing their data and all personnel contributing to obtaining the results.

## Abbreviations

COVID-19: coronavirus disease 19
I: confidence interval
ELISA: Enzyme-linked Immunosorbent Assay
IgG: immunoglobulin G
IGRA: Interferon-Gamma-Release Assay
rt-PCR: real-time polymerase chain reaction
SARS-CoV-2: Severe acute respiratory syndrome coronavirus 2

